# Clearing the fog: Is Hydroxychloroquine effective in reducing Corona virus disease-2019 progression: A randomized controlled trial

**DOI:** 10.1101/2020.07.30.20165365

**Authors:** Sultan Mehmood Kamran, Zill-e-Humayun Mirza, Arshad Naseem, Rizwan Azam, Naqeeb Ullah, Farrukh Saeed, Wasim Alamgir, Salman Saleem, Shazia Nisar, Mehmood Hussain

## Abstract

**Background:** Hydroxychloroquine (HCQ) has been considered to treat Coronavirus disease 2019 (COVID-19) but data on efficacy is conflicting. we analyzed the efficacy of HCQ) in addition to standard of care (SOC) compared with SOC alone in reducing disease progression in Mild COVID-19

**Methods:** A single centre open label randomized controlled trial during 10^th^ April to 31^st^ May 2020 was conducted at Pak emirates Military Hospital (PEMH) Five hundred patients of both genders having age between 18-80 years with Mild COVID-19 were enrolled. Patients assigned to standard dose of HCQ plus SOC were 349 while 151 patients received SOC (control group). Primary outcome was progression of disease while secondary outcome was PCR negativity on day 7 and 14. The results were analyzed on SPSS version 23. *P* value <0.05 was considered significant.

**Results:** Median age of intervention group (34 ± 11.778 years) and control group (34 ± 9.813 years). Disease progressed in 16 patients, 11 (3.15%) were in intervention group as compared to 5 (3.35%) in control group, (*P* value = 0.865). PCR negativity in intervention and control groups were (day 7, 182 (52.1%) vs. 54 (35.7%) (*P* value = 0.001), (day 14, 244 (69.9%) vs. 110 (72.8%) (*P* value = 0.508). Consecutive PCR negativity at day 7 and 14 was observed in 240 (68.8%) in intervention group compared to 108 (71.5%) in control group. (*P* value = 0.231).

**Conclusion:** Addition of HCQ to SOC in Mild COVID-19 neither stops disease progression nor help in early and sustained viral clearance.

**Clinical Trial number:** NCT04491994 available at ClinicalTrials.gov

## Introduction

Beyond supportive care, there are currently no proven treatment options for coronavirus disease (COVID-19)^1^. As mortality in patients with critical category is quite substantial^2^, hence every effort has to be made to intervene early and aggressively in order to prevent progression of disease. Globally, approximately eight million confirmed cases of Covid-19 have been reported with an outcome based overall mortality of 5.51%^3^. In Pakistan, there is exponential rise in Covid-19 cases in last few months. Nevertheless, data from various international studies shows that 81% of patients have had mild to moderate disease, which includes non-pneumonia and pneumonia cases^4^. Management of mild disease is equally important as this is the main bulk involved in transmission of disease to others. It is well known fact that asymptomatic carriers and patients with mild disease are also the main sources of disease transmissibility^5^. Therefore, it is a matter of utmost importance to detect mild cases earlier and start some investigational treatment in carefully selected hospitalized patients. Different investigational treatment options have been tried in different severity categories of COVID-19. Out of many therapeutic off-label options, HCQ seems more suitable owing to its known safety profile, side effects, posology and drug interactions^6^. HCQ has been found to have good in vitro activity against SARS-CoV-2^7^and better safety profile than chloroquine^8^. A small study on 36 patients shows that hydroxychloroquine (HCQ) treatment is significantly associated with viral load reduction/disappearance in COVID-19 patients^9^. Similarly, it has been hypothesized that HCQ might inhibit cytokine storm by reducing CD154 expression in T cells, thus reducing chances of disease progression^**10**^. Therapeutic role of HCQ can be determined by time required for virologic clearance as well as to see whether disease is getting worse or not on the basis of symptoms aggravation and monitoring laboratory markers of Cytokine release storm. In Pakistan, PEMH is the largest Covid-19 designated hospital in the country. This hospital has already treated more than 3000 Covid-19 patients so far including many asymptomatic and mild cases. On the basis of limited evidence available, HCQ was given after consent to Mild Covid-19 patients with an aim to achieve early viral clearance and prevent progression of disease. Later on, we analyzed the data to assess the response.

## Materials/subjects

This single Centre, parallel open label randomized controlled trial was carried out during 10^th^ April to 31^st^ May 2020 at department of Pulmonology, Pakistan Emirates Military Hospital (PEMH) over 500 patients from both genders between 18-80 years of age. The study design was approved by institutional ethical review committee (ERC). The study population was comprised of patients from both genders with Mild confirmed COVID-19 after their written consent. The study protocol and approval documents are available online at ClinicalTrials.gov with trial number of NCT04491994.

Sample size was calculated using OPEN EPI with 5% level of Confidence and 80% power to detect a difference and enrolment ration 2:1 between intervention and control group, at a two-sided significance level of α=0.05, of 7 days in the median time to clinical improvement between the two groups, assuming that the median time in the SOC group was 14 days and assuming 55% efficacy of HCQ in preventing disease progression and achieving viral clearance at day 7. Calculated sample size was 467, however we used a sample size of 500.

During study period, 672 confirmed PCR positive cases were assessed for eligibility. 132 did not meet selection criteria and subsequently excluded. 540 patients were then enrolled and randomized. Further 20 patients were excluded from analysis as 15 withdrew consent and 5 became symptomatic before first dose of HCQ. During follow up 13 patients were found to be deviated from advised therapy and 7 were lost to follow up yielding a final study population of 500. **Figure 1** summarizes this process. Randomization rules were designed by Dr. Wasim Alamgir together with principal investigators and implemented by an independent statistician who was not involved in data analysis. Stratified random sampling was applied to stratify all eligible patients according to age, gender and comorbidities. Computerized random number generator was used and allocation was done in 2:1 sequence. Cards with each group assignment number randomly generated by computer were placed in sequentially numbered envelopes that were opened as the patients were enrolled

**Figure 1:**
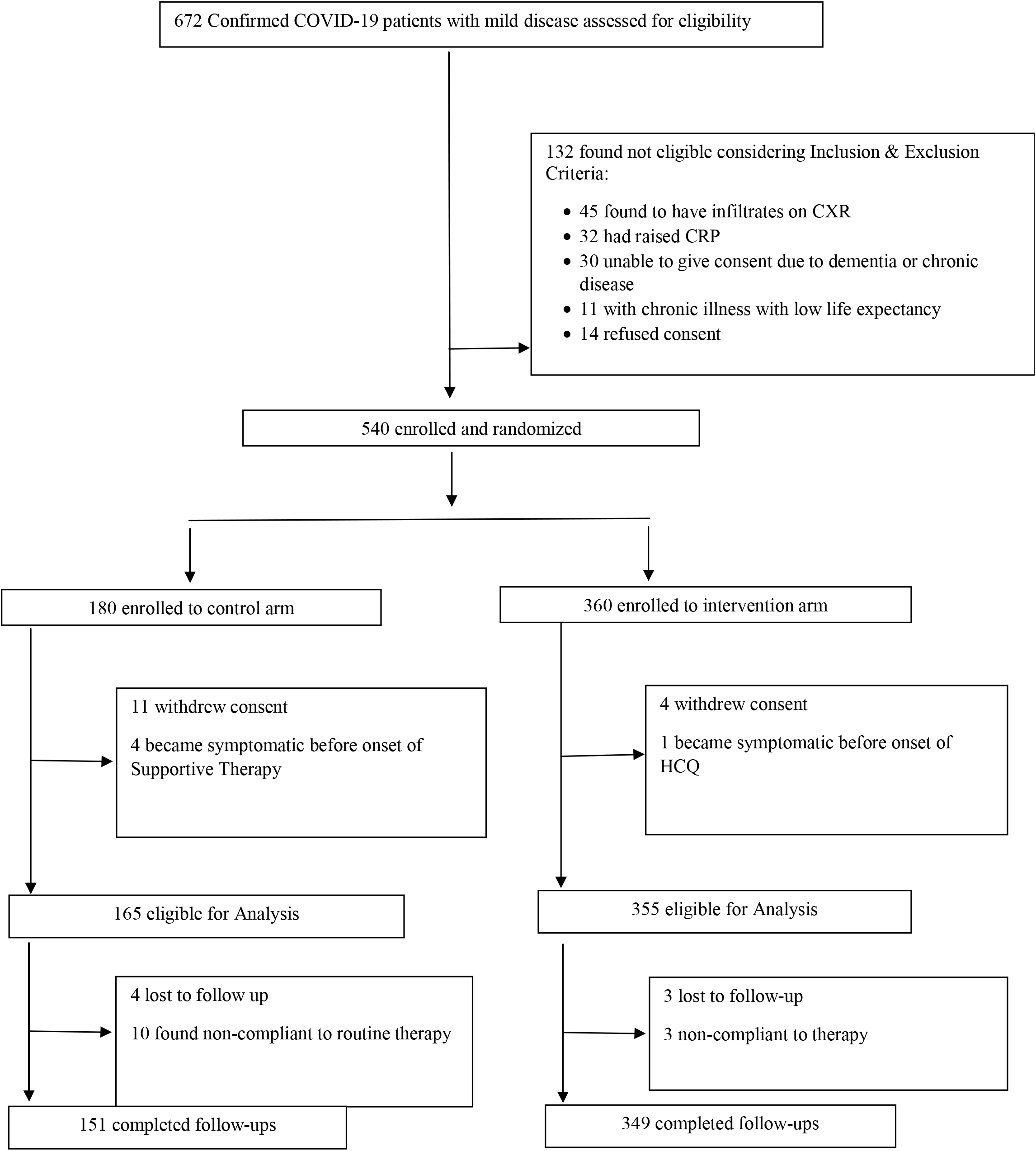
Randomization flow sheet.

A total of 349 patients included in intervention group and 151 in control group. In Hospital, HCQ was given to patients after written consent and after considering its contraindications. Three hundred and forty-nine (349) patients were included in intervention group and given HCQ in addition to SOC. After 12 hours of randomization HCQ was given. Standard dose of HCQ was 400 mg by mouth twice a day for day one followed by 200 mg 12 hourly for next 5 days. The patients who did not give consent for treatment with HCQ or had a known allergy to HCQ or chloroquine or had another known contraindication to treatment with the study drug, including retinopathy, G6PD deficiency and QT prolongation served as controls. Controls were matched with participants on the basis of age, gender and co morbids and comprised of 151 patients. Standard of care (SOC) treatment comprised of daily oral Vit C (2gms), oral Zinc (50mg), oral Vit-D (alfacalcidol 1ug) and tablet Paracetamol (for body aches/fever), intravenous fluids, hemodynamic monitoring, and laboratory testing for SARS-CoV-2 and baseline blood parameters. Neither patients, nor investigators, nor statisticians were masked to treatment assignment. Lab staff who performed sampling for PCR, basic blood tests and other routine measurement were unaware of treatment information. Side effect of drug were monitored daily. Patients on HCQ underwent daily ECG to assess QT prolongation. An increase in QT interval >25% from baseline was considered significant and HCQ stopped. Any visual complaint by the patient warrant urgent referral to eye specialist and stoppage of drug. Data regarding age, comorbidities, history of contact with a positive patient, days since contact, duration of symptoms, PCR status with date and base line labs/X-ray chest were recorded. Any patient with day 0 CRP greater than 6mg/dl, Absolute lymphocyte count (ALC) < 1000 or evidence of infiltrates on X-ray chest were excluded. Daily temperature, respiratory rate (RR) and resting O2 saturation with pulse oximetry were monitored in all patients during their hospitalization.

A case was considered confirmed on the basis of positivity of RT-PCR of combined Oropharyngeal and Nasopharyngeal swabs. Severity of disease was defined as per criteria designed by WHO^11^. Mild disease meant Patients with uncomplicated upper respiratory tract viral infection having non-specific symptoms such as low-grade fever (fever < 100F for < 3 days), fatigue, body aches, cough (with or without sputum production), anorexia, muscle pain, sore throat, nasal congestion, anosmia, headache and rarely diarrhea, nausea, and vomiting. PCR sampling was done on day 7 and 14 of admission. Any chronic health condition for which patients were on prior treatment was considered as co morbidity. After start of treatment, development of fever > 101 F for > 72 hours, shortness of breath by minimal exertion (10-Step walk test), derangement of basic lab parameters (ALC < 1000 or raised CRP) or appearance of infiltrates on CXR during course of treatment was labeled as progression irrespective of PCR status. PCR status of patients was checked after 7 days and 14 days of initiation of treatment.

Inclusion criteria included (1) Mild Corona virus disease (COVID-19) (2) PCR confirmed infection (3) Hospital admitted patients (4) 18-80 years age Exclusion criteria were (1) Moderate, severe and critical COVID-19 (2) day 0 CRP greater than 6mg/dl, ALC < 1000 or evidence of infiltrates on X-ray chest (3) comorbidity with life expectancy less than 6 months (4) Contraindications to HCQ therapy. Primary outcome was disease progression within 5 days of start of treatment. This was defined by development of fever > 101 F for > 72 hours, shortness of breath by minimal exertion (10-Step walk test), derangement of basic lab parameters (ALC < 1000 or raised CRP) or appearance of infiltrates on CXR. Patients underwent 6 hourly axillary temperature check, daily 10 feet walk test, daily Blood counts, CRP and X-Rays on day 0, 3 and 5. Secondary outcome was Viral clearance. PCR negativity on day 7 and 14 after admission was recorded. Statistical interpretation of data was performed using Statistical Package for Social Sciences (SPSS) version 23. Results were expressed as mean, standard deviation (±SD) for all continuous variables and frequency and percentage for categorical data. We used t-test and chi-square test as appropriate to the nature and distribution of the variables. A p-value < 0.05 was considered statistically significant.

## Results

During the study, a total of 500 patients of Mild COVID-19 were included, with a mean age of 35.96 ± 11.2 years (intervention group: 34 ± 11.778 vs. control group: 34 ± 9.813), Overall, males 466 (93.2%) and females 34 (6.8%) were included in trial. Male to female proportion in intervention and control groups were 328(94%) male and 21(6 %) females vs 139 (91.4%) male and 13(8.6%) females respectively. Most patients were healthy young individuals with co-morbids only in 38 (7.6%), 31(8.9%) in intervention and 7 (4.6%) in control group. Type 2 Diabetes Mellitus (DM) in 15 (3%) was the commonest disease. Positive contact history was found in 315 (63%) patients. Among constitutional symptoms, cough 163 (32.6%), low grade fever 133 (26.6%), body aches 96 (19.2%), anosmia 83(16.6%) and fatigue 56 (11.2%) were the most common. Less common symptoms were sore throat 33 (6.6%), diarrhea 21 (4.2%) and headache 21 (4.2%). Completely asymptomatic patients were 101 (20.2%). HCQ in addition to SOC treatment was given to intervention group comprising of 349 (69.8%) patients while 151 (30.2%) patients of control group received only SOC treatment.

Among 16 patients who showed disease progression **(Table-1)**, 11 (3.15%) were from intervention group, and 5 (3.3%) from control group with *P* value of 0.940. Co morbids were present in 31 (8.9%) patients in intervention group, and 7 (4.66%) (*P* value = 0.095) in control group. In intervention group, out of 11 patients with diseases progression, 4/31 (12.9%) were with co morbids as compared to 2 out of 7 (28.6%) in control group (*P* value = 0.304). Overall, Progression of disease was significantly associated with presence of co morbidities as 6 (15.8%) patients out of 38 with co morbids showed progression as compared to only 10 (2.2%) out of 462 patients without co morbids. (*P*-value < 0.00001).

**Table-1.** Assessment of Effect of HCQ on progression of disease.

| **EFFECT of HCQ** | **Treatment** |  | **p Value** |
| --- | --- | --- | --- |
|  | **HCQ plus standard of care** | **Standard of care alone** |  |
| **Overall Progression** | 11/349 (3.15%) | 5/151 (3.3%) | 0.940 |
| **Progression in co morbids** | 4/31 (12.9%) | 2/7 (28.6%) | 0.304 |

Chi-square test applied.
HCQ; Hydroxychloroquine

Overall, PCR negativity was observed in 236 (47.2%) patients on day 7 and in 354 (70.8%) patients on day 14. Effects of HCQ on PCR status of study population is given in **Table 2**. Day wise PCR negativity in intervention and control groups respectively were as follows; (day 7: 182 (52.1%) vs. 54 (35.7%) (*P* value = 0.001), (day 14; 244 (69.9%) vs. 110 (72.8%) (*P* value = 0.508). Successive day 7- and 14-day PCR negativity was observed in 240 (68.8%) patients in intervention group vs. 106 (70.1%) in control group (*P* value = 0.231) PCR remained positive in 62 (17.8%) patients of intervention group vs. 32 (21.2%) patients of control group (*P* value=0.231).

**Table-2.**
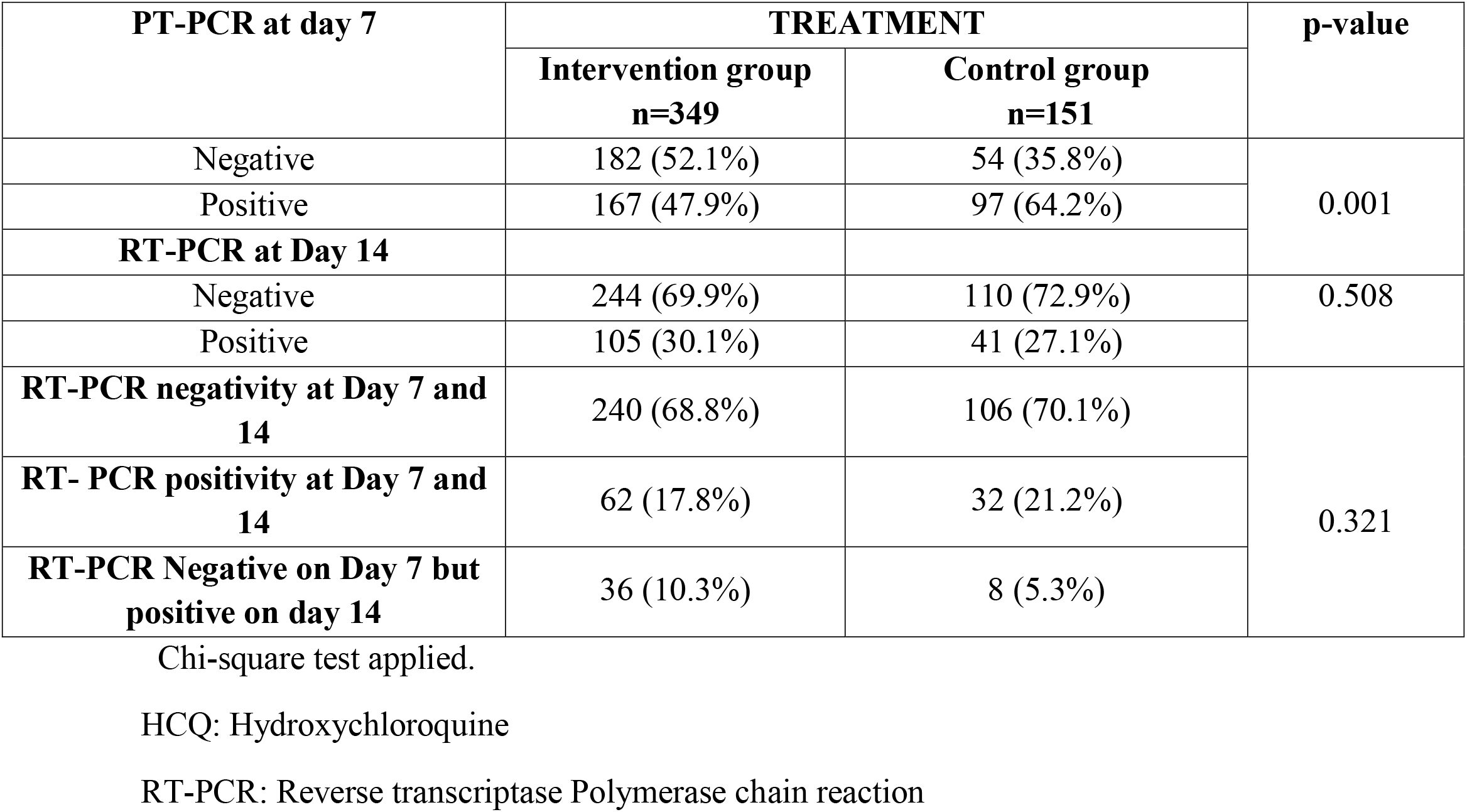
Assessment of Effect of HCQ on RT-PCR status of study population.

## Discussion

Although there was much hype about effectiveness of HCQ in treating COVID-19 but our study did not show any significant benefit of using HCQ. Firstly, HCQ did not prevent progression of disease in patients with or without co morbids although it was postulated to dampen cytokine release storm by Dan Zhou et al^**10**^. Secondly, as far as PCR negativity was concerned, its addition to supportive treatment showed significantly better early PCR negativity at day 7, but at day 14 there was not much difference in PCR negativity between the two studied groups. Nonetheless, it did not show any side effects in our study.

We used the same doses of HCQ as used by Yao X et al^**12**^ and no side effects were observed in their study also. Results of our study are also contrary to a highly publicized study done by Gao J et al^**13**^ which showed early viral clearance and decreased rate of disease progression. Comparatively it was a study with smaller sample size (n=100) and they used Chloroquine instead of HCQ used in our study. As far as viral clearance at day 7 is concerned, our results are similar to that of non-randomized control trial from France by Gautret et al^**9**^. Their study showed significantly better viral clearance at day 6 of inclusion ((70% vs. 12.5%; P = 0.001) with use of 600mg/day of HCQ for 10 days. However, in addition to HCQ, they also used Azithromycin. Although highly rated initially, this study had only 20 participants in interventional arm out of which 6 removed from study due to intolerance to medication. In addition, it was a non-randomized trial containing major biases between studied groups, and patients were not followed till day 14 to see viral clearance again. In comparison, we followed patients at day 14 and found that a subset of day 7 PCR negatives turned positive again on day 14. This observation found in our study might be because of false negative PCR at day 7 owing to variable sensitivities of testing kits or a false positive PCR at day 14 due to presence of non-infective dead viral particles. When we compare results of our study with RCT done by Chen J et al^14,^ interestingly it is found that although day 7 PCR results of our study are showing clear edge to HCQ but the primary endpoints in both studies are comparable. Chen J et al used the same dose of HCQ as in our study but in moderate COVID-19 as compared to mild category used in our research. Their study showed that HCQ did not prevent progression of disease and there was similar viral clearance between supportive treatment group and HCQ group (93.3% vs. 86.7%) (p value > 0.05) at day 7. However, sample size included 30 patients and PCR status was not checked at day 14. Our study demonstrates similar results as recommended by Infectious diseases society of America by Adarsh bhimraj et al ^**15**^. They analyzed three RCTs and six comparative cohort studies done on confirmed COVID-19 patients who were hospitalized and treated with HCQ. They studied many variables such as mortality, clinical progression, clinical improvement and adverse events and concluded that HCQ failed to show any benefit in term of viral clearance or halting progression of disease. In our study disease progression was significantly higher in patients with co morbidities even at younger age. This observation is proven in a large-scale study which had demonstrated that patients with chronic diseases are at higher risk of disease progression^**16**^. As at start of pandemic in Pakistan, our hospital had policy to admit every PCR positive case, hence, the median age of our study population was relatively younger. In our study 93.2% population was male. Overall, it has been seen that corona viruses such as severe acute respiratory syndrome (SARS)-CoV and the Middle East respiratory syndrome–CoV (MERS) predominantly affect male gender^**17**^ and may be for same genetic reasons SARS-CoV-2 is also predominantly affecting male population.

Nevertheless, there are certain limitations of our study as well. Firstly, the main subgroup in which study was done were males so the results cannot be generalized to both genders. Secondly, the study was done in mild cases and moderate/severe cases were not included so it cannot be determined whether HCQ is of any benefit in advanced COVID-19 or not. Thirdly, the patients were not followed up after discharge from the hospital hence, exact progression of disease could not be ascertained. Fourthly, we did not use quantitive RT-PCR to exactly determine the viral load which is a strong bias to affect viral clearance. Fifthly, PCR positivity at day 14 is of uncertain significance because it is now evident that after 10^th^ day onset of illness, presence of non-replicable viral nucleic acid material only, are being picked up by the PCR^**18**,**19**^ and such patients are regarded as non-infective. Finally, even with best sampling techniques, sensitivity of RT-PCR for SARS-CoV-2 ranges between 34-80%^**20**^ so exact estimation of viral clearance will definitely remain under question. Despite the limitations, our study is first of its own kind in Pakistan which is reinforced by a larger sample size and relatively longer follow up time.

## Conclusion

Our study shows that addition of HCQ to supportive treatment in mild COVID-19 cases is not significantly associated with prevention of disease progression. Despite showing significantly early PCR negativity at day 7, day 14 PCR results are similar to that of non HCQ arm. The findings of our study correlate with the results of various clinical trials done internationally.

## Data Availability

All data is available on SPSS sheet and can be shared on reasonable request

## Acknowledgement

I would like to express my deep gratitude to Professor Dr Imran Fazal, Professor Dr Zafar Ali Qureshi, Dr Kumail Abbas khan and Dr Yousaf Jamal for their patient guidance, enthusiastic encouragement and useful critiques of this research work. My grateful thanks are also extended to my colleagues; Dr Maryam Hussain, Dr Zahra, Dr Zulqernain, Dr Saeed, Dr Sher Khan and Dr Komal Arshad for their help in collecting the data.

## Disclaimer

I certify that authors have not published, posted, or submitted any related papers from the same study.

## Funding source

This research did not receive any specific grant from funding agencies in the public, commercial, or not-for-profit sectors.

I also certify that all authors have contributed significantly and that all authors are in agreement with the content of the manuscript. Many journals will refer to the definition of authorship as set up by The International Committee of Medical Journal Editors (ICMJE)

